# Teledermatology-Supported Care for Skin Neglected Tropical Diseases and Common Skin Diseases in Côte d’Ivoire: a Mixed Methods Evaluation

**DOI:** 10.64898/2026.05.11.26352967

**Authors:** Aubin Yao, Diabate Almamy, Mutiyu Akanbi Sule, Amos Sebastien Koffi, Nanan Kouassi Valentin, Kouassi Lazare Kouadio, Sakiko Itoh, Faradia Kernizan, Alyssa R. Schwinn, Ligué Agui Sylvestra Dizoé, Paul Koffi-Aboa, Mamadou Kaloga, Ronald E. Blanton, Bamba Vagamon, Rie R. Yotsu

## Abstract

**Background:** Skin-related neglected tropical diseases (skin NTDs) continue to affect people living in remote communities of endemic countries, particularly in regions with limited access to dermatological care. This operational research evaluated the impact of the eSkinHealth app, a digital health tool designed to enhance case management of skin NTDs and other skin diseases in Côte d’Ivoire. The eSkinHealth app functions as a portable electronic medical record and a platform for teledermatology, connecting frontline healthcare workers to remote specialists.

**Methodology/Principal Findings:** The study was conducted across sixteen primary health centers (PHCs) in the Sinfra and Bouaflé districts, regions endemic for skin NTDs. Using a before-and-after implementation design, baseline data were collected from paper registries and compared with data captured through the app. The primary objective was to assess changes in skin disease detection and diagnosis, while also evaluating usability, acceptability, and feasibility of the tool among healthcare workers. A total of 1,766 patients were included in the analysis (mean age 22.8 years; 55% male). During the intervention period, skin NTD registrations increased significantly from 30 to 91 cases (p < 0.01). Buruli ulcer cases rose from 6 to 14 (p = 0.05), scabies from 24 to 70 (p = 0.13), and other NTDs such as leprosy, lymphatic filariasis, and yaws were newly detected and documented. In contrast, registrations of non-NTD skin diseases decreased from 662 to 472 cases (p < 0.01); however, the proportion of non-NTD cases which received diagnostic confirmation increased markedly, from 0% at baseline to 94% during the intervention period (p < 0.01). Qualitative interviews with nurses and community health workers highlighted improvements in diagnostic accuracy, patient engagement, and confidence in daily practice, while also noting persistent challenges such as stigma, transportation barriers, technical difficulties, and patient concerns about privacy.

**Conclusions/Significance:** The integration of the eSkinHealth app into routine PHC services proved effective in enhancing diagnostic capacity for skin NTDs in resource-limited settings. However, capturing other skin diseases proved more difficult given their high prevalence. While the app demonstrated clear benefits in improving diagnostic rates and healthcare worker confidence, persistent challenges such as technical issues and patient concerns about privacy need to be addressed for future scalability. As with many digital tools, further refinement will be an ongoing process, and the lessons learnt from this study may provide valuable guidance for similar initiatives in comparable settings.

**Author Summary:** Over one billion people worldwide are affected by skin-related neglected tropical diseases (skin NTDs), yet many live in areas with limited access to dermatological care. In Cote d’Ivoire, a country with 27.5 million people, there are only 80 dermatologists, who are densely concentrated in urban centers. We developed and tested an app called eSkinHealth that allows healthcare workers in rural clinics to photograph skin conditions, record patient information, and consult with dermatologists remotely.

We implemented this tool across eight health centers across two health districts, Sinfra and Bouaflé, in central Cote d’Ivoire, that are characterized by the endemicity of numerous skin NTDs. During the seven-month intervention period, we found that cases of skin NTDs detected and documented increased three-fold compared to the previous year. Moreover, implementation of eSkinHealth supported the diagnosis and documentation of other skin conditions that had previously gone undiagnosed or unrecorded within routine care. Through interviews with nurses and community health workers, we found that the platform improved their confidence in diagnosing skin conditions and engaging with patients.

However, challenges remain, including technical difficulties, concerns about patient privacy, transportation barriers, and disease-related stigma. Our findings suggest that digital health tools can strengthen disease detection in resource-limited settings, but use of these tools can be made increasingly more effective by addressing not only technical issues, but also broader structural and social barriers to healthcare.

## Introduction

Over one billion people worldwide are at risk of, or currently affected by, neglected tropical diseases (NTDs) related to the skin, known as skin NTDs.^1^ These conditions encompass a group of diseases that manifest with signs and symptoms on the skin, involving at least ten specific diseases or disease groups as identified by the World Health Organization (WHO). In 2022, the WHO introduced a strategic framework for integrating skin NTDs, aiming to enhance care for individuals affected by skin NTDs through collaboration and resource-sharing, emphasizing broader health benefits.^2-4^

Côte d’Ivoire, a West African country, is known for its endemicity of several skin NTDs, including Buruli ulcer, leprosy, lymphatic filariasis, scabies, and yaws.^5^ We have been conducting skin surveillance to detect, diagnose, and treat these skin NTDs.^5,6^ Our findings indicate that regions with a high burden of skin NTDs often also exhibit a high incidence of other dermatological conditions. For example, one of our previous studies revealed that one in four children were affected by one or more dermatological conditions.^5^ Similar studies in different countries with skin NTD endemicity report similar challenges, highlighting the need for a holistic approach to diagnosing skin diseases and a system that provides access to dermatological care in remote areas.^7-10^ Despite the high burden, there is very limited expertise available in these places.^11^ Côte d’Ivoire, with a population of 27.5 million, has only 80 dermatologists nationwide of whom mostly reside in bigger cities.

To bridge the gap in the access to dermatological care, we have been undertaking skin surveillance initiatives with use of a digital health tool, ‘eSkinHealth’ application (app), in remote communities of Côte d’Ivoire.^12,13^ The eSkinHealth app functions as a portable electronic patient chart which offers a field-adapted platform to provide both direct diagnostic and management assistance to healthcare workers in remote settings through teledermatology.^12^ To overcome poor internet-connectivity of some areas, data entry into eSkinHealth can be done without internet connection. When users have access to internet, data can be uploaded, or ‘synchronized’, to the iCloud server, and then from this onwards can be viewed by remote personnels including dermatologists (Fig 1).

**Figure 1.**
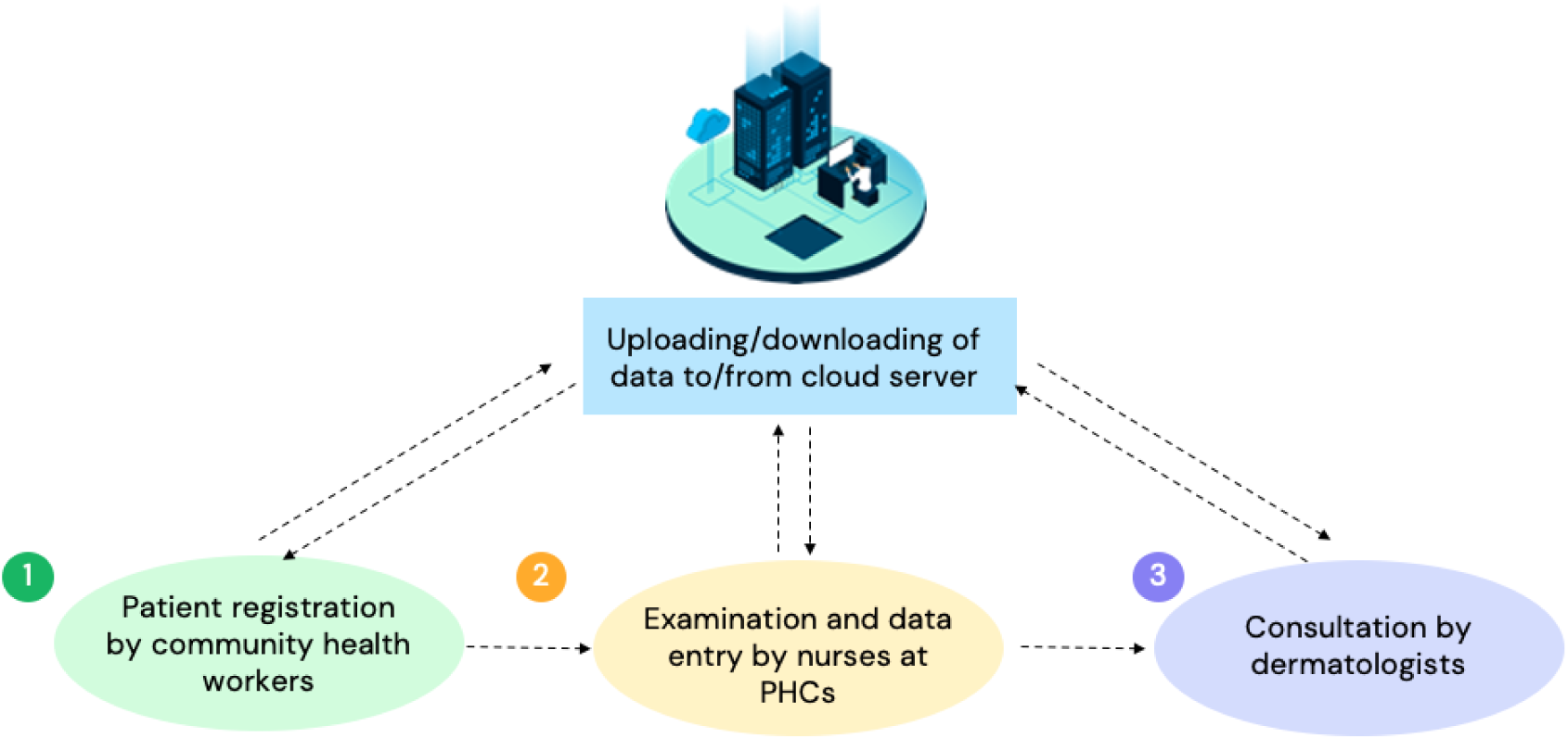
Data flow of the eSkinHealth app.

In our initial three-month pilot trial, we evaluated the usability and effectiveness of the eSkinHealth app embedded within our skin surveillance approach.^13^ The results provided strong evidence both for usability and effectiveness in improving the detection and management of skin NTDs and other skin diseases. In this extended study, we aimed to 1) compare the skin disease distribution between the pre-intervention period and period with the intervention to examine any changes that had happened, and 2) assess the usability and effectiveness of our approach, across a broader geographical area with an increased number of users.

## Methods

### Study site

Eight primary health centers (PHCs) from two districts, Sinfra and Bouaflé, known for their endemicity with multiple skin NTDs and located in central Côte d’Ivoire, were selected for the study. Selection criteria included case numbers of skin NTDs over the past three years, accessibility, geographical distribution, and population coverage.

### Study design

The study was conducted in two parts. Part one was a prospective pre-post study which included a patient population comprising individuals seeking diagnosis and treatment for their skin conditions at the designated PHCs in the two districts, who were registered in the eSkinHealth app. The patient recruitment period was from August 2022 to February 2023. Due to the evident differences in the previous three-month pilot trial, no control arm was included.^13^ Instead, we compared the intervention period against the same period of the previous year (pre-intervention, August 1, 2021 to February 28, 2022) to evaluate the prevalence of skin diseases and the intervention’s impact on diagnostic rates. Part two assessed the usability, acceptability, and feasibility of the enhanced skin surveillance approach using the eSkinHealth app among health workers.

### Part 1: Range of Skin Disease and Diagnostic Rate Study

#### Study population

Eligible patients were those presenting with any skin conditions at the selected PHCs, with no age limitations. Exclusions included individuals unable to provide consent or residing outside the targeted site.

Patient consultations and registrations in the eSkinHealth app were conducted in a private room at the PHCs. Free medications were provided for Buruli ulcer, leprosy, and yaws, supported by the National Program under the Ministry of Health. For other conditions, prescriptions were issued. In severe cases, assistance was provided for medication purchases and/or transportation fees for referrals.

#### Data collection

Patients were registered in the eSkinHealth app at their first consultation and during follow-up. The data collected in the eSkinHealth app is detailed in Supplementary File 1. Pre-intervention data, including age, sex, date of visit, clinical presentation, diagnosis, and treatment, were collected from paper-based patient records by the study staff.

#### Analysis

Statistical analyses were performed using Stata software (version 16, StataCorp) with a significance threshold of *P*<.05 in a 2-tailed test. We summarized the data by group assignment using descriptive statistics: means and SDs were used for continuous data with normal distribution, medians and IQRs for skewed data, and percentages for categorical data. As for our primary outcome, we compared the number diagnosed between control and intervention groups using the chi-square test.

### Part 2: Usability, Acceptability, and Feasibility Study

#### Study population

The study included 8 nurses and 16 community health workers (CHWs) from Sinfra District and an equal number from Bouaflé District. Participants were selected based on commitment, positive attitude, effective patient interaction, no plans to leave their post for at least one year, and technical proficiency. Participants in Sinfra District were retained from the previous study, with the exception of one nurse who was transferred.^14^ All participants were equipped with a smart tablet with the eSkinHealth app (version 1.2.8) and a Wi-Fi router. They received training on using the app by study staff and on skin diseases from dermatologists.

#### Data collection

Usability was assessed using the System Usability Scale (SUS) questionnaire at base line and the end of study at 6-months.^15,16^ Semi-structured interviews were performed at end of study to gather perceptions, opinions, and experiences regarding the project’s strengths, areas for improvement, and community impact. Interviews were performed in French (the official language of Côte d’Ivoire) and later translated into English for analysis. Duplicate responses were coded once to avoid bias. Coding was performed by ARS and reviewed and agreed by RY, with disagreements resolved by AY. The semi-structured interview guide is provided as Supplementary File 2.

#### Analysis

SUS questionnaire responses were analyzed and compared across subgroups using t-tests. Interview responses were analyzed using MAXQDA 2020 software (VERBI Software, Berlin, Germany). First, the data were thoroughly familiarized by reading and re-reading transcripts to identify initial patterns. Responses were then coded and organized according to the content of each response. Data were then mapped and interpretated within and across emerging themes. Upon conclusion of coding, responses were compared across occupational groups (CHWs vs. nurses) and health district (Sinfra vs. Bouaflé) to explore their similarities and differences between these groups to gain insights into the unique benefits, challenges, and recommendations of the intervention specific to each subgroup.

### Ethical considerations

The methodology was reviewed and approved by the institutional review board (IRB) at the Ministry of Health in Côte d’Ivoire (approval no. IRB000111917) and Tulane University (IRB 2020-2054-SPHTM). Informed consent was obtained in French, the official language of Côte d’Ivoire. For participants who spoke only a local language, a member of our study team translated the informed consent form verbally to ensure understanding. For children aged below 18, informed consent was obtained from their parents or guardians. In the case of children aged between 12 and 18 years, informed assent was also obtained. All data were de-identified to protect participant confidentiality.

## Results

### Part 1: Distribution of skin NTDs and other skin conditions

A total of 1,766 patients were included in the analysis. The mean age of the patients was 22.8 years (SD 18.3), and 55.0% were male. In the baseline group (n = 1129), the mean age was 22.6 years (SD 17.6) and 54.8% were male. In the intervention group (n = 637), the mean age was 23.2 years (SD 19.4), and 55.0% were male.

Table 1 shows the number of registered cases of skin NTDs and other skin conditions captured during the baseline period (August 2021-February 2022) using paper registries and during the intervention period (August 2022-February 2023) using the eSkinHealth app. Notably, registered Buruli ulcer cases increased from 6 to 14 (*p* = 0.05), scabies from 24 to 70 (*p* = 0.13), and other skin NTDs such as leprosy, lymphatic filariasis, and yaws – previously unrecorded – were detected and registered during the intervention period. Overall, skin NTD registrations increased significantly from 30 to 91 cases (*p* < 0.01). Conversely, the number of non-NTD skin diseases recorded decreased from 662 to 472 cases (*p* < 0.01). however, the proportion of non-NTD cases which received diagnostic confirmation increased markedly, from 0% at baseline to 94% during the intervention period (p < 0.01). The median time to complete registration and documentation of the eSkinHealth app, including photo-taking of skin lesions, was 7 minutes 16 seconds (IQR 25-75%: 4 minutes 30 seconds to 12 minutes 20 seconds).

**Table 1.** Registered case number of skin-related neglected tropical diseases (skin NTDs) and other skin conditions, baseline versus intervention.

|  | Baseline<br>(August 2021-February 2022)<sup>1</sup> |  |  | Intervention<br>(August 2022-February 2023)<sup>2</sup> |  |  | p value |
| --- | --- | --- | --- | --- | --- | --- | --- |
|  | Sinfra | Bouaflé | TOTAL | Sinfra | Bouaflé | TOTAL |  |
| *Skin NTDs* |  |  |  |  |  |  |  |
| Buruli ulcer | 3 | 3 | 6 | 9 | 5 | 14 | 0.05 |
| Leprosy | 0 | 0 | 0 | 2 | 1 | 3 | 0.36 |
| Lymphatic filariasis | 0 | 0 | 0 | 2 | 0 | 2 | 0.13 |
| Scabies | 5 | 19 | 24 | 43 | 27 | 70 | 0.13 |
| Yaws | 0 | 0 | 0 | 1 | 1 | 2 | 0.13 |
| Sub-total | 8 | 22 | 30 | 57 | 34 | 91 | <0.01 |
| *Other diseases* |  |  |  |  |  |  |  |
| Sub-total | 121 | 541 | 662 | 192 | 280 | 472 | <0.01 |
| TOTAL | 124 | 544 | 668 | 206 | 287 | 493 | <0.01 |

<sup>1</sup> Includes suspected cases only
<sup>2</sup> Includes both confirmed and suspected cases

Furthermore, Table 2 outlines the reasons for the 113 cases in the intervention group where dermatologists were unable to establish diagnoses. Of these, 16 cases (14.2%) were attributed to “synchronization error/no image” due to poor network connectivity. The majority (76 cases, 67.3%) occurred when patients failed to visit the PHC as advised by community health workers. Three cases (2.7%) were linked to poor image quality, and in 10 cases (8.9%), the images were of acceptable quality but insufficient for diagnosis based on visual assessment alone.

**Table 2.** Reasons for undiagnosed cases by dermatologists in the intervention group.

|  | Intervention |  |  |
| --- | --- | --- | --- |
|  | Sinfra | Bouaflé | TOTAL |
| Synchronization failure / no image | 4 (11.8%) | 12 (15.2%) | 16 (14.2%) |
| Only entered by CHW | 22 (64.7%) | 54 (68.4%) | 76 (67.3%) |
| Yes image but poor quality | 2 (5.9%) | 1 (1.3%) | 3 (2.7%) |
| Yes image (pure no diagnosis) | 2 (5.9%) | 8 (10.2%) | 10 (8.9%) |
| **TOTAL** | **34** | **79** | **113** |

### Part 2: Usability, Acceptability, and Feasibility Study

A total of 44 participants from the study population were interviewed as a part of the usability, acceptability, and feasibility study. In Sinfra, 21 participants were interviewed, including 8 nurses (38%) and 13 community health workers (62%). In Bouaflé, 23 participants were interviewed, including 7 nurses (30%) and 16 community health workers (70%). Table 3 summarizes participant characteristics by district. The median age was 42 years in both sites, and the majority were male (41 participants, 93.2%). All participants reported familiarity with and daily use of mobile phones or tablets. However, the frequency of computer use varied between individuals, with 25 (56.8%) reported never using a computer.

**Table 3.** Characteristics of study participants.

| Characteristics | Health District |  |  |  |
| --- | --- | --- | --- | --- |
|  | Sinfra<br>(n=21) |  | Bouaflé<br>(n=23) |  |
| **Age (years)\*** | 42 | (34–49) | 42 | (39–47) |
| <40 years | 10 | (47.6) | 9 | (39.1) |
| 40–49 years | 6 | (28.6) | 11 | (47.8) |
| >49 years | 5 | (23.8) | 3 | (13.0) |
| **Sex** |  |  |  |  |
| Male | 21 | (100.0) | 20 | (87.0) |
| Female | 0 | (0.0) | 3 | (13.0) |
| **Educational background** |  |  |  |  |
| Primary school | 6 | (28.6) | 4 | (17.4) |
| Junior high school | 5 | (23.8) | 10 | (43.5) |
| Senior high school | 2 | (9.5) | 5 | (21.7) |
| University or graduate school | 8 | (38.1) | 4 | (19.0) |
| **Occupation** |  |  |  |  |
| Community health worker | 13 | (61.9) | 16 | (69.6) |
| Nurse | 8 | (38.1) | 7 | (30.4) |
| **Working periods** |  |  |  |  |
| Less than 1 year | 3 | (14.3) | 0 | (0.0) |
| 1–5 years | 2 | (9.5) | 7 | (30.4) |
| 6–10 years | 7 | (33.3) | 7 | (30.4) |
| More than 10 years | 8 | (38.1) | 9 | (39.1) |
| **Frequency of use of cell phones/tablets** |  |  |  |  |
| Every day | 21 | (100.0) | 23 | (100.0) |
| A few times per week or less | 0 | (0.0) | 0 | (0.0) |
| **Frequency of use of computers** |  |  |  |  |
| Every day | 6 | (28.6) | 2 | (8.7) |
| A few times per week | 2 | (9.5) | 4 | (17.4) |
| A few times per month | 0 | (0.0) | 2 | (8.7) |
| A few times per year | 2 | (9.5) | 1 | (4.3) |
| Never | 11 | (52.4) | 14 | (60.9) |

\*median n (%)

### System Usability Scores (SUS) results

Table 4 presents the results of the System Usability Scores (SUS) for participants in Sinfra and Bouaflé. In Sinfra, the mean SUS score increased from 73.3 (SD 14.4) at baseline to 78.6 (SD 11.1) at the end of the study. In Bouaflé, the mean score of SUS increased more markedly, from 55.4 (SD 19.1) at baseline to 89.3 (SD 10.4) at the end of the study.

**Table 4.**
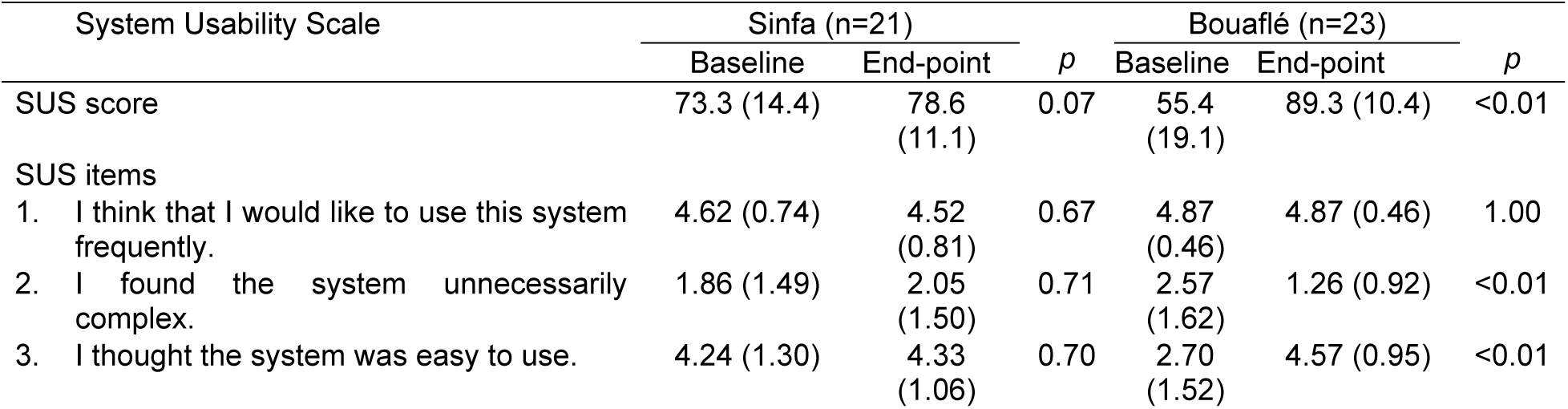
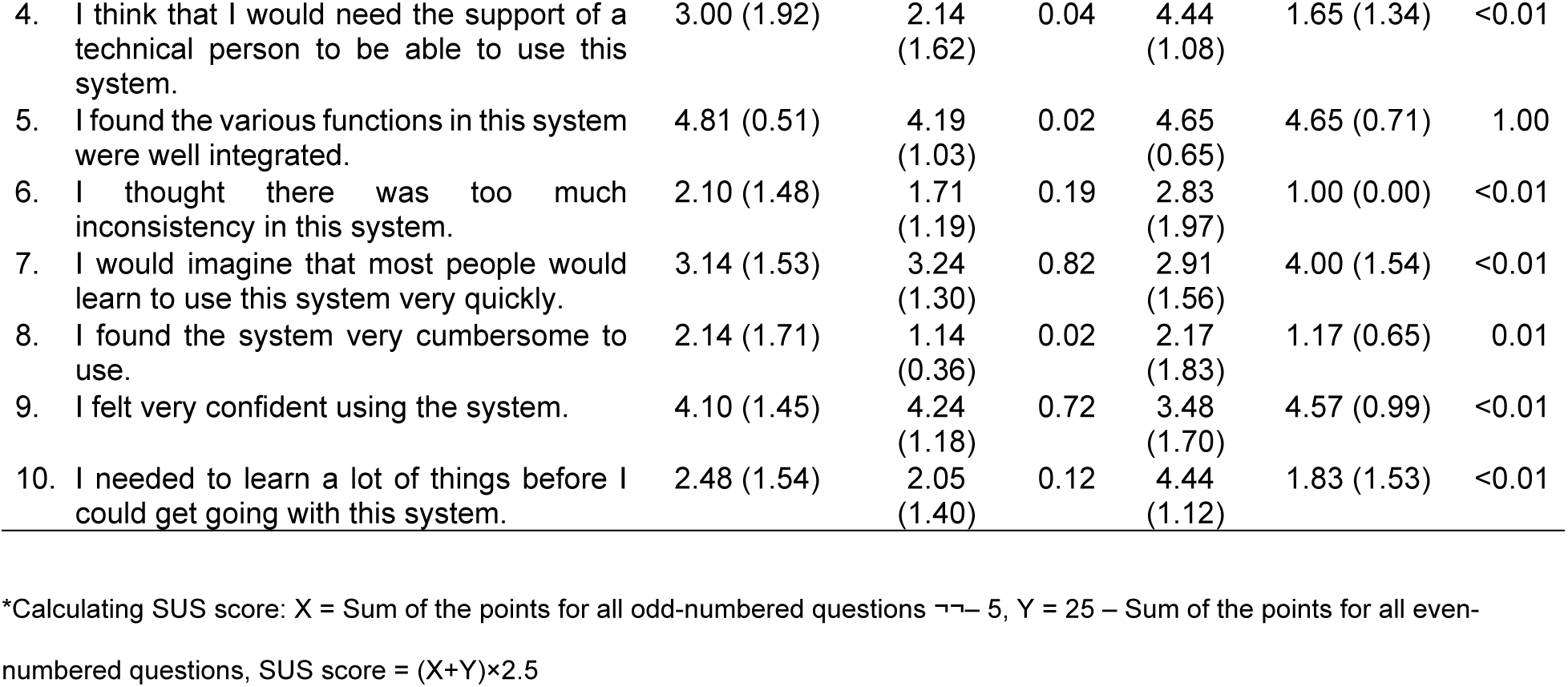
Results with system usability scores (n=44).

### Semi-structured interview results

Initial coding of the interview data identified 50 codes related to participants’ experiences with the project. Nearly all individual responses reflected one or more of these codes. These were then consolidated into 23 themes, which were further organized into five overarching categories. These categories captured participants’ perspectives on changes in clinical practice, use of digital tools, barriers to participation, access to care, and priorities for strengthening and sustaining the project, that are described in the sections below.

#### Improved practice and shifts in health-seeking behavior

Participants frequently described improved clinical practice as a major strength of the project, influencing positive changes in health seeking behavior among patients. They noted that the project enhanced the population’s awareness of the seriousness of skin NTDs, encouraging more proactive care-seeking for dermatological symptoms that had previously been ignored. Additionally, others highlighted the project’s contribution to strengthening health system capacity and active case finding efforts for dermatological conditions. Overall, the participants received the project positively and expressed a strong desire for its continuation and expansion to include other illnesses and regions.

> “It’s an opportunity for the public, because those who used to hide their illnesses now have the courage to bring them out into the open.” (CHW #1, Sinfra)

Differences emerged by occupation. CHWs across both health districts often emphasized the provision of medications and improvements in daily practice as key strengths. They valued how the project enabled more efficient and effective patient care, including treatments that were previously less accessible. Nurses, in contrast, more frequently pointed to the project’s contributions to diagnostic and surveillance improvements.

> “The project has enabled us to improve our consultations, especially for pathologies such as dermatosis, and to be more precise in our diagnoses. It has also enabled us to get closer to the population, who used to be very reluctant.” (Nurse #1, Sinfra)

#### Experiences with the tablets – from confidence and efficiency to technical challenges

Participants described both benefits and challenges related to the use of the tablet. Reported benefits included improved confidence, greater ease of diagnosis, and streamlined digital record keeping. For nurses, confidence stemmed from enhanced diagnostic capacity, while for CHWs, simply having and using the tablets gave them a stronger sense of reliability and authority in their work.

> “Yes, our practices have changed for the better, because we can now distinguish between dermatoses. To follow up patients, we don’t have to go digging through registers to find information about them.” (Nurse #3, Sinfra)
>
> “This project has made us more reliable in the eyes of the public, because the recordings are now done on the tablets.” (CHW #, Sinfra)

Challenges included initial difficulty in learning and integrating the tablet into their daily work. However, most participants noted that once they became more familiar with navigating the app, it ultimately improved their workflow. Location-specific issues also emerged. In Bouaflé, some participants reported skepticism from the population about using the tablet in medical care, whereas in Sinfra, technological unreliability was more commonly described, particularly difficulties with synchronization (uploading and downloading data from the server). Internet connectivity was also frequently raised as a barrier.

#### Perceived barriers to participation - stigma, privacy, and unmet expectations

Nurses and CHWs reported that some patients refused to register due to feelings of shame or stigma associated with their condition, or concerns about the privacy of their photos, especially when lesions were in private areas. In some cases, parents refused to allow their children’s photos to be taken. These concerns were raised more often in Bouaflé than in Sinfra.

> “…this [refusal] case was a woman and the disease was in her genital area, and given the tablet, she was worried that everyone would be able to see her photo. But I explained to her that it’s only the superior who sees in order to prescribe the treatment.” (Nurse #1, Bouaflé)

Some participants mentioned that the population believed medication would be provided as part of the project. This expectation sometimes led to disappointment or reluctance when medicines were not available. People expected free medications, as they were accustomed to during mass drug administration (MDA) campaigns for other conditions.

> “The lack of medication for diagnostic cases often poses a problem, as the population thinks that medication is free.” (Nurse #1, Bouaflé)
>
> “After the prescription, they never bought the drugs because they have no means.” (Nurse #1, Bouaflé)

#### Access to Care – digital tools easing but not eliminating transportation barriers

Participants frequently described transportation as the major factor shaping access to care. Some noted that the project acted as a facilitator by enhancing access to care through remote support, reducing the need for patients to travel to health centers or hospitals - ‘reducing distance’- and helping them ‘save transportation costs’.

> “The photo from the CHWs goes directly to the doctors, as some patients find it difficult to go to the health center.” (CHW #2, Bouaflé)

At the same time, long distances, difficult terrain especially during rainy seasons, and lack of proper means of transport continued to pose challenges. These barriers affected both patients to travel for consultations or follow-up visits or for CHWs to visit them at their homes.

> “No [patient did not refuse]. We register them, but the problem is getting them to the health centers.” (CHW #1, Bouaflé)
>
> “…Once patients have been registered by the CHWs, it is difficult for them to come to the health centers.” (Nurse #2, Bouaflé)

#### Strengthening and sustaining the project – scope, support, and awareness

Participants made several recommendations to improve and sustain the project. Four main themes emerged: expanding the scope beyond the current target diseases, increasing the number of trained personnel, ensuring project regularity and consistency, and raising overall awareness of the project in the population.

##### Expanding scope of training

Nurses frequently recommended expanding training to cover a broader range of diseases beyond the skin NTDs and selected common skin conditions currently included, emphasizing that more conditions should be addressed. Participants also highlighted the need for training on the use of tablets so that all PHC staff could work at the same level.

> “Train all health center staff, allocate tablets so that everyone has the same level of training and information, and send tablets that can take chips [SIM cards].” (Nurse #1, Bouaflé)
>
> “The diseases most often encountered are not those treated by the project [*i.e*., skin NTDs].” (CHW #3, Bouaflé)

##### Project consistency

Participants across both districts emphasized the importance of consistency, including regular monitoring by the local team and timely payments to CHWs.

> “For the last three months, the CHWs have been working without pay, they’re discouraged.” (Nurse #2, Sinfra)

##### Support for outreach and access

Several CHWs recommended additional material support to help them overcome persistent transport barriers. Requests included rain protection, backpacks for carrying equipment, and motorbikes to reach remote areas. These suggestions directly reflected the challenges they faced in providing care to patients who lived far from health centers or in difficult-to-access locations.

##### Raising awareness

Finally, participants stressed the need to increase community awareness of the project, particularly in encouraging people to visit PHCs and register. Active case-finding and direct outreach were seen as essential to overcome initial reluctance.

> “Yes, first with the population. It was difficult to get them to come to the health centers and show their disease, so we had to do a lot of awareness-raising to convince them. Also, through active case-finding activities.” (Nurse #1, Sinfra)

## Discussions

This mixed-methods study evaluated the implementation of the eSkinHealth app in primary health care settings in Côte d’Ivoire, with a focus on its role in case detection and diagnosis of skin-related conditions, as well as its usability, acceptability and feasibility. As digital health tools gain more prominence across various disease control programs, this study explored how such technologies can support in improving access to dermatological care in underserved communities – not only for skin NTDs but for skin diseases more broadly. Building on skin NTD integration framework,^2-4^ the World Health Assembly adopted the Resolution on “Skin Diseases as a Global Public Health Priority” (WHA 78.15) in May 2025.^17^ One of the priority actions listed under this resolution is “to consider innovative, integrated service delivery models, including telemedicine platforms and training for digital assessments, to strengthen dermatology services, especially in remote and hard-to-reach areas.”^17^ Our findings contribute directly to this global agenda by demonstrating both the opportunities and challenges of using digital tools such as eSkinHealth to operationalize integrated skin health services at the community level.

Overall, findings indicate that the platform offered meaningful opportunities to strengthen frontline clinical practice, enhance diagnostic capacity for skin NTDs, and positively influence health-seeking behavior within underserved communities. Healthcare providers described improvements in confidence, workflow efficiency, and engagement with patients, while increased awareness of skin diseases appeared to encourage earlier and more proactive care-seeking for conditions that had previously been neglected or hidden. The platform enhanced diagnostic capacity for skin NTDs, especially with increased detection of Buruli ulcer and scabies.

However, while the total number of diagnosed skin diseases increased, the number of registered overall cases was lower than the baseline, possibly reflecting the additional perceived workload associated with digital registration. The digitalization of healthcare offers clear benefits but is also known to increase administration burden, which may contribute to user fatigue and burnout.^18-20^ The median time to complete registration – including documentation and photography – was 7 minutes 16 seconds, a duration that is generally acceptable in clinical workflows but may still deter busy healthcare providers.^21^ Although the app includes simplified process for follow-up visits, this function was not frequently used, largely because many patients did not return to the clinic for follow-up care.

A major implementation challenge arose from incomplete data uploads due to internet connectivity limitations. Such infrastructural weaknesses have been widely reported in mHealth interventions across Africa.^22,23^ Although eSkinHealth was designed with off-line functionality, infrastructural constraints still critically impacted its intended functionality and hindered the smooth communication of patient information among healthcare providers. Large image file sizes and unstable connections disrupted data transmission, limiting the teledermatology performance, as also reported in the literature.^24^

Building a robust off-line electronic medical record system is technically challenging because, unlike 24/7 online systems, offline platforms must reconcile data stored locally on multiple devices, manage synchronization queues, handle conflicts between versions, and ensure files to be uploaded successfully once connectivity is available.^25,26^ These requirements add layers of technical complexity, increasing the risk of incomplete uploads, data loss, or delays in information flow. Although mHealth tools with offline functionality, such as the eSkinHealth app, offer important advantages in low-connectivity environments and emergency situations, the specific challenges associated with off-line systems and strategies to overcome them remain insufficiently documented, largely because only a limited number of digital health tools currently incorporate such features. ^25,26^ This gap highlights the need for further research to better understand how offline-capable systems can be optimized and scaled.

The quantitative usability and qualitative findings provided complementary insights into the eSkinHealth user experience, a key factor in guiding evidence-based improvements to the app’s functionality and value for its target users. Overall, the platform was well received by the participants, many of whom highlighted its benefits to clinical practice, including enhanced diagnostic accuracy and improved community trust in healthcare providers. Similar findings were reported in the 2020 pilot evaluation of LeishCare® in Brazil, where healthcare professionals frequently noted that the app facilitated communication among providers and improved diagnostic capacity and recognition of leishmaniasis as a differential diagnosis in the region.^27^

The SUS demonstrated substantial improvement over the study period, indicating a shift from “marginal” to “excellent” usability.^15,16^ Item-level analysis showed substantial increases in perceived ease of use, reduction in perceived complexity, and decreased need for technical support over time. These quantitative findings were reinforced by interview data, in which participants reported growing confidence in using the system and described it as less cumbersome, reflecting increased self-efficacy and improved workflow integration. Such findings are consistent with previous mHealth evaluations in low-resource settings, which have documented rapid usability gains after hands-on practice and peer learning.^28-30^ Moreover, the very high endpoint SUS scores (above 80) correspond to high user satisfaction and suggest a strong likelihood of sustained adoption if infrastructural and motivational factors are adequately supported.

When stratified by district, SUS improvement was more pronounced in Bouaflé (from 55.4 to 89.3) than in Sinfra (from 73.3 to 78.6). This difference suggests that participants with lower initial familiarity with the app or digital devices experienced greater relative gains after structured training and repeated field use. It also indicates that usability barriers are surmountable through continuous engagement, technical support, and experience-based learning rather than being intrinsic to the technology itself. Conversely, the smaller increment in Sinfra - where users had prior pilot experience - may reflect a ceiling effect, with usability already at a satisfactory level.^31^

Despite these gains, insufficient community awareness was repeatedly cited as a barrier. Although behavioral changes were observed among registered patients, this suggested that much of the broader population was still unaware of the services provided through the app, highlighting an implementation gap that could undermine long-term sustainability.^32^ Beyond community mobilization, participants identified persistent challenges such as poor internet connectivity and transportation limitations. Intermittent internet connectivity posed a barrier to the integration of the eSkinHealth application, as described earlier. Insufficient transportation has long been recognized as a major barrier to healthcare access among vulnerable populations in rural areas and a significant demotivating factor among CHWs worldwide.^33-35^ While digital health tools may help mitigate some of these obstacles, they cannot fully resolve logistical constraints, nor are they applicable in all clinical contexts.^36^

In contrast, concerns related to privacy and stigma emerged specifically in relation to the use of digital health tools. The expansion of mHealth technologies in recent decades has introduced new considerations influencing healthcare accessibility, including the need to protect patient data, promote digital literacy, and ensure reliable mHealth infrastructure.^37-40^ In our setting, concerns regarding privacy and stigma were frequently raised and are consistent with reports from other digital health interventions. For example, in a recent mobile phone–based tuberculosis treatment support program in Uganda, patients feared community exposure, particularly among women.^41^ Addressing these sensitivities requires not only technological safeguards but also community-based engagement and trust-building strategies.

This study had several limitations. Two rural districts with comparable settings were our study sites which may limit generalizability to other context with differing health system structures and infrastructures. Additionally, in many cases, diagnoses were made remotely, and a true “gold standard” face-to-face dermatological assessment was not feasible. The qualitative component included only healthcare providers, excluding patients and community members; thus, the study may not fully capture the perspectives of service beneficiaries. In our future studies, we plan to include patients, their families, and community members to strengthen participatory evaluation – and to ensure for sustainability.

Telemedicine and digital innovations are increasingly recognized as promising approaches to strengthening health service delivery, particularly in settings where specialist availability is limited.^42^ Framing digital health tools, including the eSkinHealth app, as complementary tools to the existing healthcare infrastructure may offer a more effective and sustainable approach to promoting equitable dermatological care across diverse social and economic contexts. While there are clear benefits – such as increasing the number of patients diagnosed, successful implementation require careful attention to health-system readiness, infrastructure, and user experience. Ultimately, the continued development of eSkinHealth platform is intended to support more equitable and accessible dermatological care in underserved settings and contribute to the shared goal of skin health for all.

## Data Availability

All relevant data are provided within the manuscript and its supporting information files. The data collected in the eSkinHealth application is provided as supporting information (S1 File). The script used to guide semi-structured interviews is also provided as supporting information (S2 File).

## Acknowledgements

We are grateful to the district officers, nurses, and community healthcare workers of Sinfra and Bouaflé Health Districts of Côte d’Ivoire for their contributions in this project. We also would like to thank all the patients who took part in this study.

## Supporting Information

**Table S1.** Data list in the eSkinHealth app.

**Table S2.** Semi-structured interview guide. Interviewers were permitted to ask clarifying follow-up questions to each primary question if needed.

## References

1. Yotsu RR, Fuller LC, Murdoch ME, et al. World Health Organization strategic framework for integrated control and management of skin-related neglected tropical diseases: what does this mean for dermatologists? Br J Dermatol 2022.

2. World Health Organization. Ending the neglect to attain the Sustainable Development Goals: a strategic framework for integrated control and management of skin-related neglected tropical diseases. Geneva, Switzerland, 2022.

3. Yotsu RR, Fuller LC, Murdoch I, et al. A global call for action to tackle skin-related neglected tropical diseases (skin NTDs) through integration: an ambitious step change. PLoS Negl Trop Dis 2023.

4. Yotsu RR, Fuller LC, Murdoch ME, et al. World Health Organization strategic framework for integrated control and management of skin-related neglected tropical diseases: what does this mean for dermatologists? Br J Dermatol 2023; 188(2): 157–9.

5. Yotsu RR, Kouadio K, Vagamon B, et al. Skin disease prevalence study in schoolchildren in rural Cote d’Ivoire: Implications for integration of neglected skin diseases (skin NTDs). PLoS Negl Trop Dis 2018; 12(5): e0006489.

6. Yotsu RR, Comoe CC, Ainyakou GT, et al. Impact of common skin diseases on children in rural Cote d’Ivoire with leprosy and Buruli ulcer co-endemicity: A mixed methods study. PLoS Negl Trop Dis 2020; 14(5): e0008291.

7. Saka B, Kassang P, Gnossike P, et al. Prevalence of skin Neglected Tropical Diseases and superficial fungal infections in two peri-urban schools and one rural community setting in Togo. PLoS Negl Trop Dis 2022; 16(12): e0010697.

8. Gnimavo RS, Fajloun F, Al-Bayssari C, et al. Importance of consultations using mobile teams in the screening and treatment of neglected tropical skin diseases in Benin. PLoS Negl Trop Dis 2023; 17(5): e0011314.

9. Hogewoning A, Amoah A, Bavinck JN, et al. Skin diseases among schoolchildren in Ghana, Gabon, and Rwanda. Int J Dermatol 2013; 52(5): 589–600.

10. Komba EV, Mgonda YM. The spectrum of dermatological disorders among primary school children in Dar es Salaam. BMC Public Health 2010; 10: 765.

11. Mosam A, Todd G. Dermatology Training in Africa: Successes and Challenges. Dermatologic clinics 2021; 39(1): 57–71.

12. Yotsu RR, Itoh S, Yao KA, et al. The Early Detection and Case Management of Skin Diseases With an mHealth App (eSkinHealth): Protocol for a Mixed Methods Pilot Study in Cote d’Ivoire. JMIR Res Protoc 2022; 11(9): e39867.

13. Yotsu RR, Almamy D, Vagamon B, et al. An mHealth App (eSkinHealth) for Detecting and Managing Skin Diseases in Resource-Limited Settings: Mixed Methods Pilot Study. JMIR Dermatol 2023; (6): e46295.

14. Yotsu RR, Almamy D, Vagamon B, et al. An mHealth App (eSkinHealth) for Detecting and Managing Skin Diseases in Resource-Limited Settings: Mixed Methods Pilot Study. JMIR Dermatol 2023; 6: e46295.

15. Brooke J. SUS: A “quick and dirty” usability scale. London: Taylor and Francis; 1996.

16. Brooke J. SUS: a retrospective. Journal of Usability Studies 2013; 8(2): 29–40.

17. World Health Assembly. Skin diseases as a global public health priority. In: Assembly WH, editor. WHA7815. Geneva, Switzerland; 2025.

18. Wosny M, Strasser LM, Hastings J. Experience of Health Care Professionals Using Digital Tools in the Hospital: Qualitative Systematic Review. JMIR Hum Factors 2023; 10: e50357.

19. Arslan F, Marcus J, Khatami A, Guergachi A, Keshavjee K. Towards a Regulatory Framework for Workflow Improvement in Electronic Medical Records. Stud Health Technol Inform 2024; 312: 54–8.

20. Nongrum MS, Dhaliwal BK, Na Y, et al. Disconnected data: mHealth data systems and challenges for primary health care workers in India. SSM-Health Systems 2025; 5.

21. Irving G, Neves AL, Dambha-Miller H, et al. International variations in primary care physician consultation time: a systematic review of 67 countries. BMJ Open 2017; 7(10): e017902.

22. Mycetoma due to Nocardia. Arch Dermatol 1970; 102(3): 345–6.

23. Olu O, Muneene D, Bataringaya JE, et al. How Can Digital Health Technologies Contribute to Sustainable Attainment of Universal Health Coverage in Africa? A Perspective. Front Public Health 2019; 7: 341.

24. Coates SJ, Kvedar J, Granstein RD. Teledermatology: from historical perspective to emerging techniques of the modern era: part II: Emerging technologies in teledermatology, limitations and future directions. J Am Acad Dermatol 2015; 72(4): 577–86; quiz 87–8.

25. Ashworth H, Ebrahim S, Ebrahim H, Bhaiwala Z, Chilazi M., A Free Open-Source, Offline Digital Health System for Refugee Care. JMIR Med Inform 2022; 10(2): e33848.

26. Shrestha A, Alawa J, Ashworth H, Essar MY. Innovation is needed in creating electronic health records for humanitarian crises and displaced populations. Front Digit Health 2022; 4: 939168.

27. Ferreira da Silva PE, Junior G, Ambrozio RB, et al. LeishCare((R)): A Software Designed for the Management of Individuals with Leishmaniases. Am J Trop Med Hyg 2020; 103(2): 909–16.

28. Ide N, Hardy V, Chirambo G, et al. People Welcomed This Innovation with Two Hands: A Qualitative Report of an mHealth Intervention for Community Case Management in Malawi. Ann Glob Health 2019; 85(1).

29. Odendaal WA, Anstey Watkins J, Leon N, et al. Health workers’ perceptions and experiences of using mHealth technologies to deliver primary healthcare services: a qualitative evidence synthesis. Cochrane Database Syst Rev 2020; 3(3): CD011942.

30. Zaidi S, Kazi AM, Riaz A, et al. Operability, Usefulness, and Task-Technology Fit of an mHealth App for Delivering Primary Health Care Services by Community Health Workers in Underserved Areas of Pakistan and Afghanistan: Qualitative Study. J Med Internet Res 2020; 22(9): e18414.

31. Itoh S, Kouadio K, Didier KY, Ugai K, Yao KA, Yotsu RR. Evaluation of the Usability of a Mobile Application on Neglected Skin Diseases in Cote d’Ivoire: A Pilot Study. Stud Health Technol Inform 2022; 290: 972–6.

32. Musoke D, Ndejjo R, Atusingwize E, Mukama T, Ssemugabo C, Gibson L. Performance of community health workers and associated factors in a rural community in Wakiso district, Uganda. Afr Health Sci 2019; 19(3): 2784–97.

33. Kok MC, Muula AS. Motivation and job satisfaction of health surveillance assistants in Mwanza, Malawi: an explorative study. Malawi Med J 2013; 25(1): 5–11.

34. Kok MC, Dieleman M, Taegtmeyer M, et al. Which intervention design factors influence performance of community health workers in low- and middle-income countries? A systematic review. Health Policy Plan 2015; 30(9): 1207–27.

35. Takasugi T, Lee AC. Why do community health workers volunteer? A qualitative study in Kenya. Public Health 2012; 126(10): 839–45.

36. Kaburi BB, Wyss K, Kenu E, et al. Facilitators and Barriers in the Implementation of a Digital Surveillance and Outbreak Response System in Ghana Before and During the COVID-19 Pandemic: Qualitative Analysis of Stakeholder Interviews. JMIR Form Res 2023; 7: e45715.

37. Yew SQ, Trivedi D, Adanan NIH, Chew BH. Facilitators and Barriers to the Implementation of Digital Health Technologies in Hospital Settings in Lower- and Middle-Income Countries Since the Onset of the COVID-19 Pandemic: Scoping Review. J Med Internet Res 2025; 27: e63482.

38. Aovare P, Beune E, Laar A, Moens N, Moll van Charante EP, Agyemang C. User experiences with a mobile health app for self-management of diabetes and hypertension in Ghana: a qualitative study. Ann Med 2025; 57(1): 2517395.

39. Keddy KH, Saha S, Kariuki S, et al. Using big data and mobile health to manage diarrhoeal disease in children in low-income and middle-income countries: societal barriers and ethical implications. Lancet Infect Dis 2022; 22(5): e130–e42.

40. Hackett KM, Kazemi M, Sellen DW. Keeping secrets in the cloud: Mobile phones, data security and privacy within the context of pregnancy and childbirth in Tanzania. Soc Sci Med 2018; 211: 190–7.

41. Leddy A, Ggita J, Berger CA, et al. Barriers and Facilitators to Implementing a Digital Adherence Technology for Tuberculosis Treatment Supervision in Uganda: Qualitative Study. J Med Internet Res 2023; 25: e38828.

42. Craig A, Lawford H, Miller M, et al. Use of Technology to Support Health Care Providers Delivering Care in Low- and Lower-Middle-Income Countries: Systematic Umbrella Review. J Med Internet Res 2025; 27: e66288.

